# Description and performance evaluation of two diet quality scores based on the Nova classification system

**DOI:** 10.1101/2023.05.19.23290255

**Authors:** Caroline dos Santos Costa, Francine Silva dos Santos, Kamila Tiemann Gabe, Eurídice Martinez Steele, Fernanda Helena Marrocos Leite, Neha Khandpur, Fernanda Rauber, Maria Laura da Costa Louzada, Renata Bertazzi Levy, Carlos Augusto Monteiro

## Abstract

**Background and objectives:** The consumption of unprocessed or minimally processed whole plant foods and of ultra-processed foods, as defined by the Nova food classification system, are associated in opposite ways with diet quality and risk of diseases. However, it can be difficult to evaluate and monitor the consumption of these foods in some contexts due to lack of resources and time constraints for data collection. This study aimed to describe two simple and easily derived diet quality scores and evaluate their performance in reflecting the dietary share of unprocessed or minimally processed whole plant foods and ultra-processed foods.

**Methods:** A total of 812 adults (18 years old or older) answered the Nova24h screener, a 2-minute self-administered questionnaire that measures the consumption of a set of foods on the day before the interview. Food items included in this tool belong to two main groups of Nova classification: unprocessed or minimally processed whole plant foods (WPF, 33 food items) and ultra-processed foods (UPF, 23 food items). Two scores are obtained from this tool by summing the number of items checked - the Nova-WPF and the Nova-UPF. We compared the Nova-WPF and the Nova-UPF scores with the dietary intake (% of total energy) of all unprocessed or minimally processed whole plant foods and all ultra-processed foods, respectively, obtained through a full self-administered web-based 24-hour recall, applied on the same day. We evaluated the relationship between the approximate quintiles or intervals of each score and the corresponding % of energy intake by linear regression, and the agreement between the intervals of each score with the intervals of the corresponding % of energy intake, using the Prevalence-Adjusted and Bias-Adjusted Kappa (PABAK).

**Results:** Approximate quintiles of each score presented a direct and linear relationship with the corresponding % of energy intake (p-value for linear trend <0.001). We found a substantial agreement between the intervals of each score and of the corresponding % of energy intake (PABAK 0.72, 95% CI 0.64-0.81 for the Nova-WPF score and PABAK 0.79, 95% CI 0.69-0.88, for the Nova-UPF score).

**Conclusions:** These two scores performed well against the dietary share of unprocessed or minimally processed whole plant foods and ultra-processed foods in Brazil and can thus be used to evaluate and monitor diet quality.

## Introduction

Dietary patterns that prioritize the consumption of unprocessed or minimally processed whole plant foods – encompassing a diverse combination of fruits, vegetables, legumes, and whole grains while limiting the intake of animal-source foods – along with the avoidance of ultra-processed foods (UPFs), have been recommended to improve both human and planetary health^1-4^. There is a large body of evidence on the benefits of a higher consumption of unprocessed or minimally processed whole plant foods to the nutritional profile of diets, human health, and the environment sustainability^2, 3, 5^. At the same time, the dietary share of UPFs has been consistently associated with overall deterioration of the nutrient profile of diets^6, 7^ and a higher risk of several non-communicable diseases (NCDs)^8-11^.

Both the above-mentioned least and most processed food groups belong to two of the four groups of the Nova classification, a framework that categorizes foods and beverages according to the extent and purpose of industrial processing they have undergone^1^. The first group comprises unprocessed and minimally processed foods, which are subjected to processes that largely preserve the food matrix and do not involve the addition of salt, sugar, fat, or any other food substance^1^. Therefore, whole plant foods fall into this Nova group. The most processed group, on the other hand, is made up of ultra-processed foods, which are industrial formulations consisting mostly of substances derived from foods and cosmetic additives (i.e. flavor enhancers, colors, emulsifiers, sweeteners, and thickeners)^1^.

Despite the recommendations for healthy and sustainable diets, global evidence from repeated national food consumption and sales surveys suggests a shift away from traditional dietary patterns towards those based on animal-sourced foods and UPFs^12-19^. Thus, evaluating and monitoring food consumption to track these shifts in dietary patterns around the world seems essential. However, conventional comprehensive tools, such as 24h-recalls or food frequency questionnaires, are not designed to collect dietary data aligned with food processing and require personnel and time resources that are not always available in low-and middle-income countries.

To address part of these gaps, the current study presents, for the first time, the Nova24h screener, a novel, short and practical 24-hour recall screener that was developed to easily assess the dietary intakes of two critical Nova groups in Brazil – the unprocessed or minimally processed whole plant foods and the ultra-processed foods. Scores reflecting the level of these intakes can also be calculated from this screener. The screener consists of two lists of yes/no questions regarding the consumption of the most commonly consumed foods in Brazil, as determined by a nationally representative survey: one containing commonly consumed unprocessed or minimally processed whole plant foods grouped into 33 questions, and the other containing ultra-processed foods grouped into 23 questions^20^.

Previous research has presented the ultra-processed food’s list of the Nova24h screener and tested the Nova-UPF score in a small convenience sample of one city of the southeast region of Brazil, indicating that it has the potential to reflect the dietary share of ultra-processed foods^21^. However, the performance of the score for the unprocessed/ minimally processed whole plant food component has not yet been evaluated nor the performance of the two scores in larger samples and across sociodemographic subgroups. Thus, this study aims to (1) describe two simple diet quality scores based on the Nova classification system, calculated from the Nova24h screener, and (2) evaluate their ability to reflect the dietary share of unprocessed or minimally processed whole plant foods and ultra-processed foods, using data from a large Brazilian study.

## Methods

### Sample selection

This is a cross-sectional study undertaken with participants of the NutriNet-Brasil study, an ongoing web-based cohort that currently includes over 100,000 adult volunteers (18 years old or older) from all the regions of Brazil. Recruitment started on January 26, 2020 and is largely based on multimedia campaigns. This cohort aims to prospectively study the association between dietary patterns and chronic diseases. After study registration in the project website (https://nutrinetbrasil.fsp.usp.br/) and agreeing to participate, participants are asked to answer brief questionnaires about diet, health status, socioeconomic conditions, and other determinants of health every three months in the same website using a cellphone, tablet, or computer. The ethics committee of the School of Public Health from São Paulo University (process No. 88455417.8.0000.5421) approved the study.

For the current study, we invited a random subsample of NutriNet-Brasil participants (n 1,800) to answer on the same day, firstly the Nova24h screener, from which two diet quality scores are obtained, and then a full 24-hour dietary recall, which allows to estimate the dietary intake (% of total energy) of all unprocessed or minimally processed whole plant foods and all ultra-processed foods. Invitations were balanced across regions of the country and sex (n = 180 invitations per strata) and the data collection was carried out over a two-month period (September and October 2020).

### Data collection

The Nova24h screener, a 2-minute self-administered questionnaire, asks participants about the intake of two lists of foods consumed on the day before (checkbox format, yes/no). The first list includes varieties of Nova unprocessed or minimally processed foods that are whole plant foods (grouped into 33 items) and the second, varieties of ultra-processed foods including both plant-based and animal-based products (grouped into 23 items). For both groups, the items included in the screener represent the most consumed in Brazil according to the national food consumption survey conducted by the 2008–2009 Household Budget Survey (POF) of the Brazilian Institute of Geography and Statistics^20^.

Unprocessed or minimally processed whole plant food items are grouped in six categories: fruits, excluding fruit juices (10 items); leafy vegetables (9 items); other vegetables, excluding roots and tubers as potato and manioc (9 items); whole grains (3 items); legumes (1 item); and nuts (1 item), while ultra-processed foods items are grouped in three categories: beverages (6 items); ready-to-eat products created to replace meals (10 items); and products often consumed as snacks (7 items). The complete list of foods included in the Nova24h screener can be seen in **Supplementary material**.

Two scores are obtained from the Nova24h screener, based on the simple sum of the checked items within each food group -the Nova score of whole plant foods (Nova-WPF, ranging from zero to 33), and the Nova score of ultra-processed foods (the Nova-UPF score, ranging from zero to 23) (Supplementary material). To evaluate the performance of the Nova-WPF and the Nova-UPF scores against the % of total energy from all unprocessed or minimally processed whole plant foods (excluding roots and tubers) and all ultra-processed foods, respectively, we used data collected through a full validated self-administered web-based 24-hour recall, specifically designed to capture the consumption of each of the four Nova food groups (hereinafter called Nova24h)^22^. In the Nova24h, participants inform all foods and drinks they had consumed on the day before and their respective amount, based on a predetermined list of foods and standardized categories. Nova24h was previously validated against a standard interviewer-led multiple pass 24-hour dietary recall. Description and validation details are explained in a previously published paper^22^.

Briefly, the food list integrated into the Nova24h was developed using nationally representative food consumption data and the food grouping structure of the Brazilian Household Budget Survey 2008-2009^20^. Detailed information is also requested regarding food source, preparation method, and additions (other foods or culinary ingredients added to preparations). All food items and recipes (the latter previously disaggregated into individual ingredients) are linked with the Brazilian Table of Food Composition 7.0 (TBCA) as the primary source, or with the United States Department of Agriculture (USDA) National Nutrient Database^23, 24^ to obtain energy, macronutrient, and micronutrient contents. Each item is also classified, using a standard method, according to the extent and purpose of industrial food processing established by the Nova system into four groups: unprocessed or minimally processed foods, processed culinary ingredients, processed foods, and ultra-processed foods.

For the comparison with the Nova-WPF score, we calculated the percentage (%) of energy contribution from unprocessed or minimally processed whole plant foods, including all fruits, vegetables, whole grains, legumes, and nuts reported by the participants in the Nova24h; for the comparison with the Nova-UPF score, we calculated the percentage (%) of energy contribution from all ultra-processed foods reported in the Nova24h.

Sociodemographic information was collected in the recruitment of the participants, including geographic region of the country (North, Northeast, Southeast, South, Mid-West), sex (male, female), schooling level (0-11, 12+, in years), and age range (18-34, 35-59, 60+, in years).

### Statistical analysis

First, we described the prevalence (%) of consumption on the day before the interview of each food item included in the Nova24h screener and the distribution of the Nova-WPF and Nova-UPF scores. Then, we presented the mean dietary share (% of total energy intake with 95% CI) of the unprocessed and minimally processed whole plant foods and of ultra-processed foods obtained from the full 24-hour recall, according to the intervals of each score (0-4, 5-6, 7, 8-9, 10+ for the Nova-WPF score; and 0, 1, 2, 3, 4+ for the Nova-UPF score), using linear regression models to analyze linear trend.

Lastly, we evaluated the agreement between approximate quintiles of the scores and of the respective reference measures. For that, we firstly intended to generate quintiles for the Nova scores but, considering the nature of repetitions in scores variables, the distribution of participants did not correspond to 20% in each generated category. Thus, hereinafter we refer to these categories as “approximate” quintiles or intervals. Considering the unequal distribution of participants across intervals of the scores, we chose to replicate the marginal proportions of these intervals to categorize participants according to their 24-hour recall dietary share of whole plant foods or ultra-processed foods, in order to obtain comparable groups. In this way, the distribution of the dietary share of unprocessed or minimally processed whole plant foods was divided into five parts by applying the same proportions of the Nova-WPF score approximate quintiles. The same was done for the dietary share of ultra-processed foods based on the proportions of the Nova-UPF score approximate quintiles. For example, if 30% of the participants were part of the first category of a score, we would replicate this proportion for the first interval of the reference measure. After that, we compared individuals’ classification according to the Nova-WPF score intervals with the classification according to intervals of the dietary share (% of the total energy) of whole plant foods. Likewise, we compared individuals’ classification according to the Nova-UPF score intervals with the one according to intervals of the dietary share (% of the total energy) of UPF. We calculated the PABAK (Prevalence-Adjusted and Bias-Adjusted Kappa) to estimate the agreement between the classifications based on the Nova-WPF or Nova-UPF scores with the ones based on the percentage of total energy from whole plant foods or ultra-processed foods, respectively. This index is a modification of the kappa statistic that adjusts for prevalence and bias. While kappa is highly sensitive to the prevalence of the condition, PABAK depends only on the observed agreement and is particularly useful when data are imbalanced, where one category is more prevalent than others^25^. All the agreement analyses were replicated for each socioeconomic strata (geographic region, sex, schooling level, and age range) to confirm the performance in these population subgroups. Values greater than 0.80 indicate an almost perfect agreement; between 0.61 and 0.80, a substantial agreement; between 0.41 and 0.60, moderate; between 0.21 and 0.40, fair; and equal to or less than 0.20, slight^26^.

All analyses were run using the Stata® statistical package, version 16.1 (StataCorp. 2019. Stata Statistical Software: Release 16. College Station, TX: StataCorp LLC), except for the agreement analyses, which were performed in RStudio using the statistical package “irrCAC”, where the 95% confidence intervals were also calculated through the matrix’s square weights^27^.

## Results

A total of 812 participants from the NutriNet-Brasil answered the Nova24h screener and were included in the analysis of this study. As planned, they were similarly distributed across gender and the five Brazilian regions. Participants included young (18-34 y, 36.6%), middle-age (35-59 y, 51.6%) and older (60y+, 11.8%) adults. Similarly to what is observed in the total sample of the Nutrinet-Brasil, most participants (85.6%) had university education (**Supplementary material**).

Among the unprocessed or minimally processed whole plant foods included in the Nova-WPF score, tomato, beans/lentils/chickpeas, banana, lettuce, and carrot were the five most frequently consumed items. Each of these foods were consumed by 40% or more of participants on the previous day. Conversely, bread, chocolate bar, soda, reconstituted meat products, and margarine, each of which were consumed by 15% to 20% of participants on the day before, were the most frequently consumed ultra-processed items (**Figure 1**).

**Figure 1.**
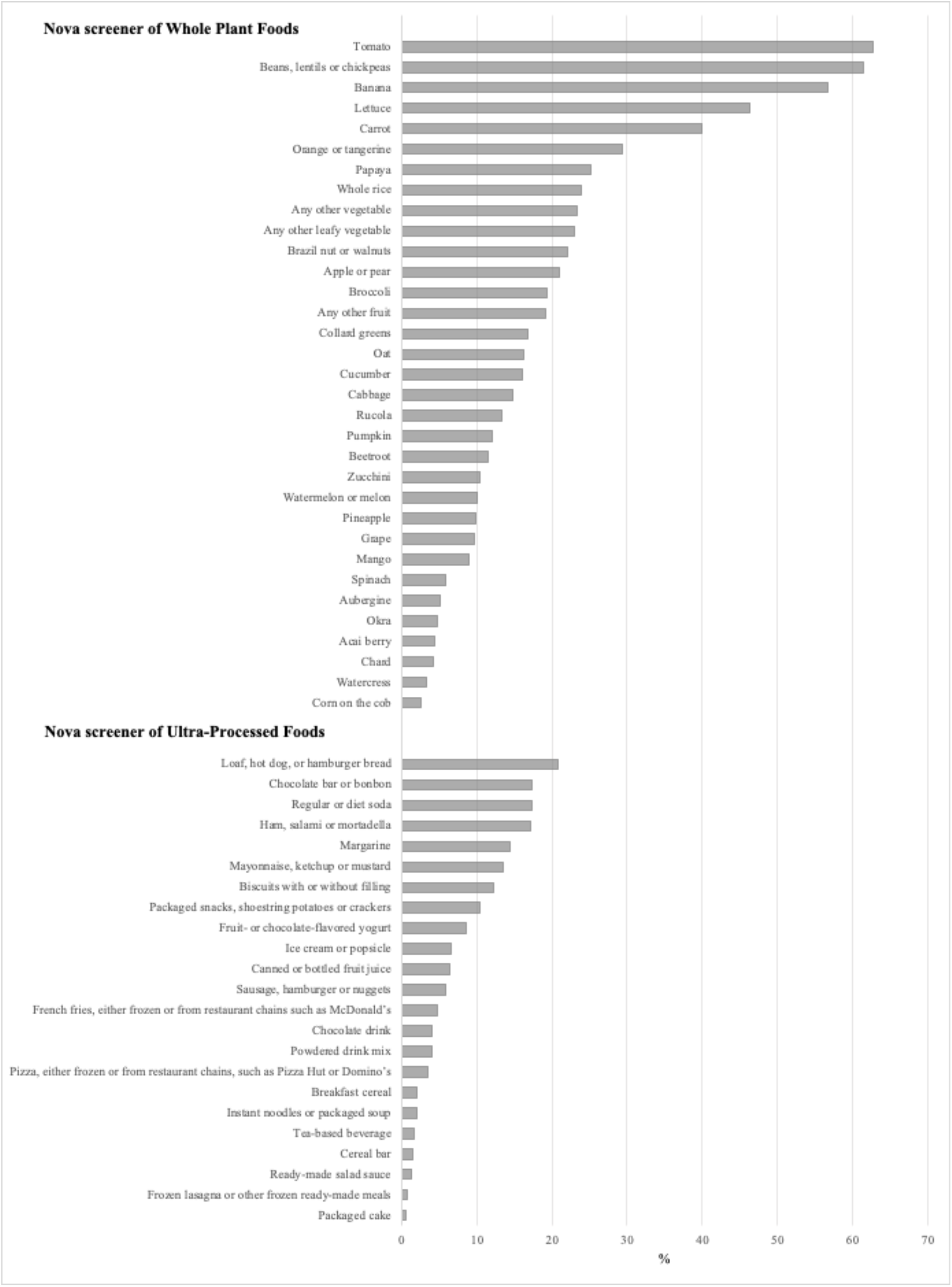
Proportion (%) of consumption on the day before of the food items included in the Nova24h screener. Participants (n 812) of the NutriNet-Brasil cohort (2020).

The Nova-WPF score presented a nearly normal distribution, ranging from zero to 20 food items consumed on the day before. Almost one fifty of the participants, 18.1%, scored 10+ (highest interval). The Nova-UPF score distribution ranged from zero to 15 and was right-skewed. Almost one fourth of the participants (23.5%) scored null, 30.2% scored one and 20.8% scored two, while 13.7% scored 4+ (highest interval) (**Figure 2**).

**Figure 2.**
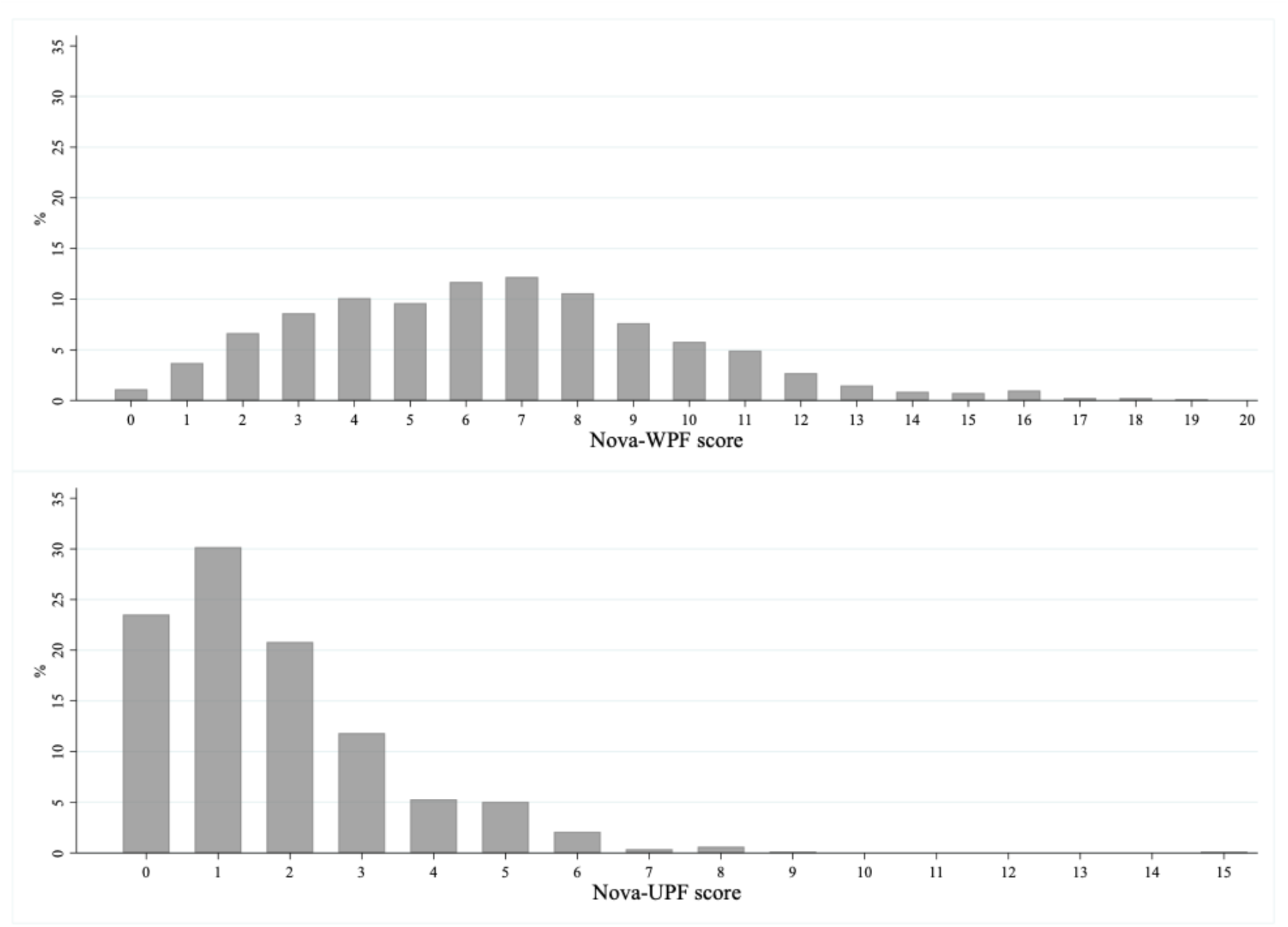
Distribution of the Nova-WPF score and the Nova-UPF score. Participants (n 812) of the NutriNet-Brasil cohort (2020). The Nova-WPF score (Nova score of Whole Plant Foods) ranges from zero to 33 and the Nova-UPF score (Nova score of Ultra-Processed Foods), from zero to 23.

Figure 3. shows that the dietary share of all unprocessed or minimally processed whole plant foods (% of total energy intake from these foods, estimated by the full 24-hour recall) increased linearly with the increase in the intervals of the Nova-WPF score (p-value for linear trend <0.001), while the dietary share of ultra-processed foods (% of total energy intake from these foods, estimated by the full 24-hour recall) increased linearly with the increase in the intervals of the Nova-UPF score (p-value for linear trend <0.001).

**Figure 3.**
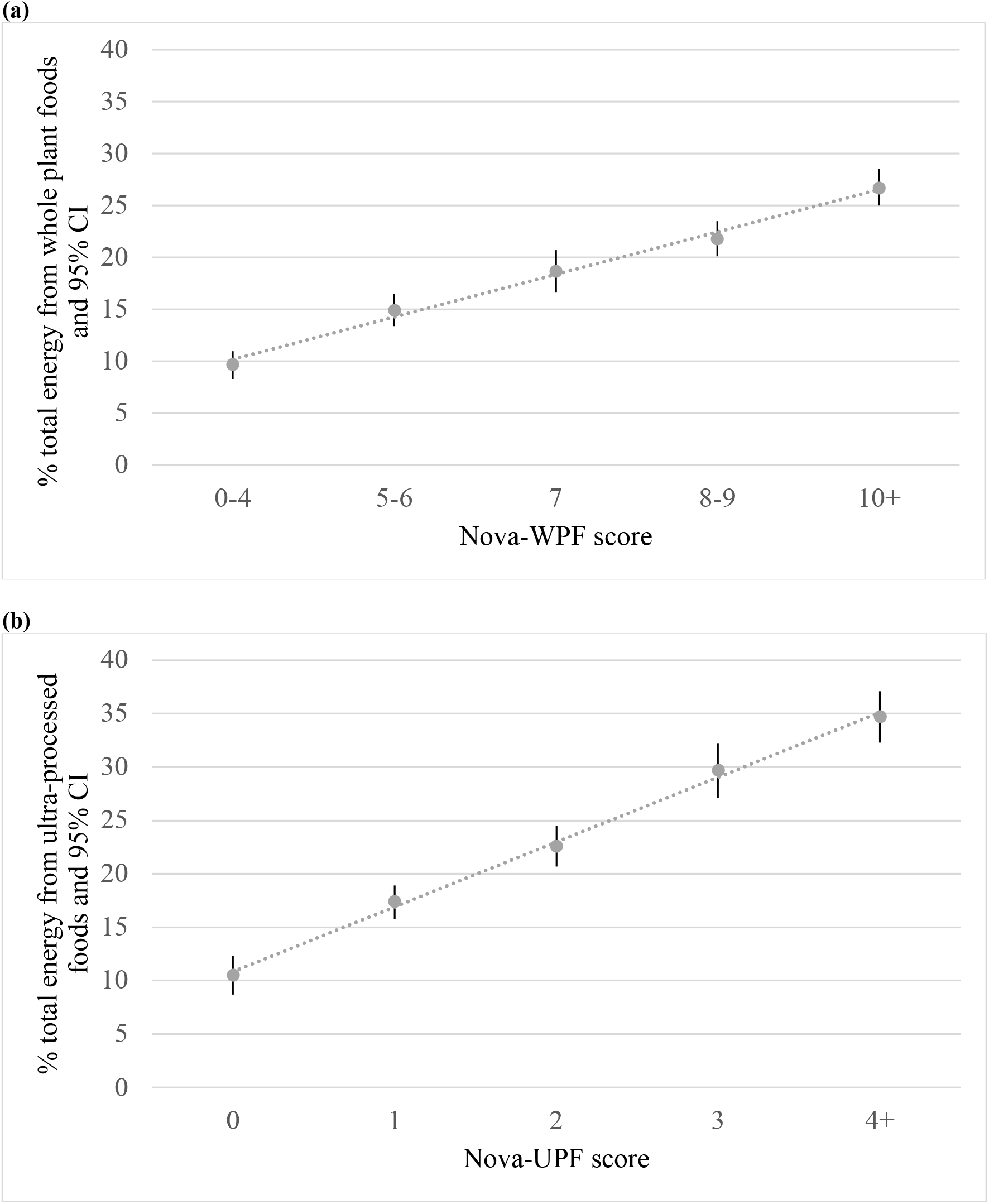
Mean dietary share (% of total energy intake with 95% CI) of (a) whole plant foods obtained from the full 24-h recall according to Nova-WPF score intervals and (b) ultra-processed foods obtained from the full 24-h recall according to Nova-UPF score intervals. Participants (n 812) of the NutriNet-Brasil cohort (2020). Nova-WPF score, Nova score of whole plant foods; Nova-UPF score, Nova score of ultra-processed foods. p-value for linear trend <0.001 in all associations.

There was a substantial agreement between the distribution of participants according to intervals of the Nova-WPF score and corresponding intervals of the dietary share of unprocessed or minimally processed whole plant foods (PABAK of 0.72; 95% CI 0.64-0.81). A substantial agreement was also observed between intervals of the Nova-UPF score and corresponding intervals of the dietary share of ultra-processed foods (PABAK of 0.79; 95% CI 0.69-0.88) (**Tables 1 and 2**). Similar agreements were observed in all sociodemographic strata (geographic regions of the country, sex, schooling level, and age range; **Table S1**).

**Table 1.** Agreement between participants classification according to Nova-WPF score intervals, estimated by the Nova24h screener, and corresponding intervals of the dietary share of whole plant foods, estimated using a full 24h dietary recall. Participants (n 812) of the NutriNet-Brasil cohort (2020).

| % of total energy intake<br>from whole plant foods<br>(full 24h recall) | Nova-WPF score intervals (Nova24h screener) |  |  |  |  |  | PABAK (95% CI) |
| --- | --- | --- | --- | --- | --- | --- | --- |
|  | 0-4 | 5-6 | 7 | 8-9 | 10+ | Total |  |
|  |  |  |  |  |  |  | 0.72 (0.64 - 0.81) |
| ≤9.67 | 17.2 | 7.6 | 1.8 | 2.0 | 1.5 | 30.1 |  |
| 9.68-15.35 | 6.8 | 4.8 | 3.6 | 4.1 | 2.1 | 21.4 |  |
| 15.36-19.34 | 2.3 | 3.1 | 1.8 | 2.7 | 2.2 | 12.1 |  |
| 19.35-27.40 | 2.8 | 3.1 | 3.2 | 4.8 | 4.3 | 18.2 |  |
| ≥27.41 | 1.0 | 2.7 | 1.7 | 4.7 | 8.0 | 18.1 |  |
| **Total** | 30.1 | 21.3 | 12.1 | 18.3 | 18.1 | 100.0 |  |

PABAK, Prevalence-Adjusted and Bias-Adjusted Kappa; Nova-WPF score, Nova score of whole plant foods.

**Table 2.** Agreement between participants classification according to Nova-UPF score intervals, estimated by the Nova24h screener, and corresponding intervals of the dietary share of ultra-processed foods, estimated using a full 24h dietary recall. Participants (n 812) of the NutriNet-Brasil cohort (2020).

| % of total energy intake<br>from ultra-processed<br>foods (full 24h recall) | Nova-UPF score intervals (Nova24h screener) |  |  |  |  |  | PABAK (95% CI) |
| --- | --- | --- | --- | --- | --- | --- | --- |
|  | 0 | 1 | 2 | 3 | 4+ | Total |  |
|  |  |  |  |  |  |  | 0.79 (0.69 - 0.88) |
| ≤8.77 | 12.6 | 8.3 | 2.0 | 0.4 | 0.4 | 23.7 |  |
| 8.78-19.55 | 7.1 | 10.5 | 8.1 | 2.5 | 2.1 | 30.3 |  |
| 19.56-28.37 | 2.2 | 5.7 | 5.2 | 4.4 | 3.2 | 20.7 |  |
| 28.38-37.85 | 1.0 | 3.4 | 3.6 | 1.8 | 2.0 | 11.8 |  |
| ≥37.86 | 0.6 | 2.3 | 2.0 | 2.7 | 6.0 | 13.6 |  |
| **Total** | 23.5 | 30.2 | 20.9 | 11.8 | 13.7 | 100.0 |  |

PABAK, Prevalence-Adjusted and Bias-Adjusted Kappa; Nova-UPF score, Nova score of ultra-processed foods.

## Discussion

This study aimed to describe two diet quality scores based on the Nova classification system and to evaluate their performance in reflecting the dietary share of unprocessed or minimally processed whole plant foods and ultra-processed foods. Both Nova scores of unprocessed or minimally processed whole plant foods and of ultra-processed foods, assessed with a simple 2-minute screener, presented significant direct and linear relationships with the dietary share (% of total energy intake) of these foods assessed with a validated self-administered web-based 24-hour recall. Substantial agreement was found between participants’ classification according to the intervals of each score and corresponding intervals of the dietary share of the foods included in each score.

A previous study evaluated, in a convenience sample of 300 individuals from São Paulo city, the performance of the Nova-UPF score in reflecting the % of total energy from ultra-processed foods, also finding a substantial agreement with the reference measure^21^. Our study adds to the field by confirming this performance in a larger and more diverse sample, and across categories of geographic regions, sex, schooling level, and age groups. This is also the first study to evaluate the performance of the unprocessed or minimally processed whole plant foods score in reflecting the dietary share of unprocessed or minimally processed whole plant foods.

Regarding the Nova-WPF score, it is noteworthy that the first group of Nova classification refers to a range of unprocessed or minimally processed foods – including animal-sourced unprocessed or minimally processed foods. However, although the intake of this whole Nova group is associated with a better dietary nutritional profile^5^, we included in the score only the food groups that have been consistently pointed out as protective against NCDs^2, 3, 28^. Also, the scores as proposed in the present study are in consonance with the current dietary recommendations, including the Planetary Health Diet and the Dietary Guidelines for the Brazilian Population, since they allow to monitor and evaluate the consumption of foods that are recommended to prefer and to avoid or limit^2, 29^.

Although we found substantial agreement in ranking individuals according to the intervals of the % of total energy from both food groups, it is important to highlight that the agreement between the scores and the reference measure was highest in the extreme intervals. The low variability of the scores might have led to a reduced accuracy in classifying individuals in the intermediate categories since a classification change can occur even with an increase or decrease in only one point of the score and the probability of classification bias in intermediate categories is higher than in extreme categories. Particularly for the Nova-UPF score, most of the population scored only up to 3 of the 23 points, which is consistent with the findings of previous studies using the same or a similar score^21, 30, 31^. However, both the low consumers according to the Nova-WPF score (those with the lowest consumption of unprocessed or minimally processed whole plant foods) and the high consumers according to the Nova-UPF score (those with the highest consumption of ultra-processed foods) ranked well, precisely those that represent the most critical population subgroups when aiming to provide information to policymakers. Moreover, we found a dose-response relationship between the scores’ intervals and the reference measure, as well as a substantial agreement in ranking individuals according to the intervals of the % of total energy from both food groups, which suggests a good discriminatory power when used as a continuous measurement.

We are aware that our study has limitations. Our sample was mostly based on participants with a high schooling level likely because the study is fully carried out through the internet and the questionnaires are self-reported. However, a review of food consumption validation studies recommended a sample size of at least 50 to 100 subjects for each stratum when evaluating population subgroups, which was achieved even for low schooled participants. Also, the scores and the reference measure could be affected by the same source of error since they are both self-reported and the recall bias or the intentional underreporting of some food items are possible for both scores under evaluation and the reference measures. Considering this scenario, it would be relevant to validate the performance of the scores in predicting health outcomes in future studies.

These two scores that (a) capture the most consumed unprocessed or minimally processed whole plant foods and of ultra-processed foods, (b) present a good performance against 24-hour recall measures of food consumption based on the Nova classification system, and (c) are calculated using data from a low research burden screener (low cost, rapid application, and straightforward to analyze), can facilitate measuring and monitoring the quality of diets, and the risk of related NCDs. The dominance of ultra-processed foods in the current food environment at the expense of unprocessed or minimally processed whole foods and the likely related increase in risk of NCDs such as obesity, cancer, diabetes, hypertension, cardiovascular diseases, and depression, as well as all-cause mortality, contribute to the relevance of the Nova24h screener and its scores, allowing for the collection of information that can orient policies and actions in public health. Finally, even though the Nova24h screener and its scores were developed based on the Brazilian context, they can be easily adapted to be used in or validated for other countries (as currently happening in Ecuador, India and Senegal)^32^, using data from national dietary intake or purchase surveys and/or other methodologies to check for the appropriateness of the screener items and for the selection of context-specific examples within each item. The screener presents potential to be used in or adapted for other countries, representing a possible tool to be applied in monitoring and evaluation systems around the world.

## Data Availability

All data produced in the present study are available upon reasonable request to the authors

## Acknowledgements

This work was funded through the São Paulo Research Foundation, FAPESP (Grant No. 2021/10993-3) and the Innovative Methods and Metrics for Agriculture and Nutrition Action (IMMANA) programme (Grant No. IMMANA 3.06), led by the London School of Hygiene & Tropical Medicine (LSHTM), which does not necessarily share its positions. FR is a beneficiary of a research fellowship from the World Cancer Research Fund.

