## Supplementary material for "Description and performance evaluation of two diet quality scores based on the Nova classification system"

**Nova24h screener**

Please take a few minutes to recall all the **foods and drinks** you consumed **YESTERDAY**, from the moment you woke up to the time you went to sleep.

| See this list of fruits and check all the ones you ate YESTERDAY: | Score |
| --- | --- |
| □ Banana | 01 |
| □ Orange or tangerine | 01 |
| □ Mango | 01 |
| □ Papaya | 01 |
| □ Pineapple | 01 |
| □ Watermelon or melon | 01 |
| □ Apple or pear | 01 |
| □ Grape | 01 |
| □ Acai berry | 01 |
| □ Any other fruit | 01 |
| □ I did not eat any fruit yesterday | - |
| See this list of leafy vegetables and check all the ones you ate YESTERDAY: |  |
| □ Lettuce | 01 |
| □ Chard | 01 |
| □ Watercress | 01 |
| □ Rucola | 01 |
| □ Collard greens | 01 |
| □ Cabbage | 01 |
| □ Broccoli | 01 |
| □ Spinach | 01 |
| □ Any other leafy vegetable | 01 |
| □ I did not eat any leafy vegetable yesterday | - |
| See this list of other vegetables and check all the ones you ate YESTERDAY: |  |
| □ Tomato | 01 |
| □ Cucumber | 01 |
| □ Carrot | 01 |
| □ Beetroot | 01 |
| □ Pumpkin | 01 |
| □ Zucchini | 01 |
| □ Aubergine | 01 |
| □ Okra | 01 |
| □ Any other vegetable | 01 |
| □ I did not eat any vegetable yesterday | - |
| See this list of foods and check all the ones you ate **YESTERDAY:** |  |
| □ Beans, lentils or chickpeas | 01 |
| □ Whole rice | 01 |
| □ Oat | 01 |
| □ Corn on the cob | 01 |
| □ Brazil nut or walnuts | 01 |
| □ I did not eat any of the foods on this list yesterday | - |
| **Total score for unprocessed or minimally processed foods** | **33** |
| See this list of drinks and check all the ones you drank **YESTERDAY:** | Score |
| □ Regular or diet soda | 01 |
| □ Canned or bottled fruit juice (Del Valle-type) | 01 |
| □ Powdered drink mix (Tang-type) | 01 |
| □ Chocolate drink (Nescau-type) | 01 |
| □ Tea-based beverage (Ice tea-type) | 01 |
| □ Fruit- or chocolate-flavored yogurt | 01 |
| □ I did not drink any of the drinks on this list yesterday | - |
| See this list of foods and check all the ones you ate **YESTERDAY:** |  |
| □ Sausage, hamburger or nuggets | 01 |
| □ Ham, salami or mortadella | 01 |
| □ Loaf, hot dog or hamburger bread | 01 |
| □ Margarine | 01 |
| □ Mayonnaise, ketchup or mustard | 01 |
| □ Ready-made salad sauce | 01 |
| □ French fries, either frozen or from restaurant chains such as McDonald’s | 01 |
| □ Pizza, either frozen or from restaurant chains, such as Pizza Hut or Domino’s | 01 |
| □ Instant noodles (Miojo-type) or packaged soup | 01 |
| □ Frozen lasagna or other frozen ready-made meals | 01 |
| □ I did not eat any of the foods on this list yesterday | - |
| See this other list of foods and check all the ones you ate **YESTERDAY:** |  |
| □ Packaged snacks, shoestring potatoes or crackers | 01 |
| □ Biscuits with or without filling | 01 |
| □ Packaged cake | 01 |
| □ Cereal bar | 01 |
| □ Ice cream or popsicle | 01 |
| □ Chocolate bar or bonbon | 01 |
| □ Breakfast cereal (Sucrilhos-type) | 01 |
| □ I did not eat any of the foods on this list yesterday | - |
| **Total score for ultra-processed foods** | **23** |

**Table S1.** Sample distribution according to sociodemographic strata and agreement between participants classification according to Nova-WPF and Nova-UPF scores intervals, estimated by the Nova24h screener, and intervals of the dietary share of whole plant foods and ultra-processed foods, respectively, estimated using a full 24h dietary recall, for each stratum. Participants (n 812) of the NutriNet-Brasil cohort (2020).

| **Variable** | **Sample** | **PABAK (95% CI)** | |
| --- | --- | --- | --- |
|  | n (%) | **Nova-WPF score** | **Nova-UPF score** |
| Geographic region |  |  |  |
| North | 123 (15.2) | 0.70 (0.47-0.93) | 0.79 (0.57-1.00) |
| Northeast | 161 (19.8) | 0.73 (0.53-0.94) | 0.77 (0.58-0.97) |
| Southeast | 179 (22.0) | 0.71 (0.51-0.90) | 0.75 (0.54-0.95) |
| South | 185 (22.8) | 0.74 (0.56-0.93) | 0.73 (0.53-0.92) |
| Mid-West | 164 (20.2) | 0.74 (0.56-0.93) | 0.79 (0.56-1.00) |
| Sex |  |  |  |
| Male | 384 (47.3) | 0.71 (0.58-0.85) | 0.75 (0.61-0.88) |
| Female | 428 (52.7) | 0.74 (0.62-0.86) | 0.77 (0.65-0.90) |
| Schooling level (y) |  |  |  |
| <12 | 117 (14.4) | 0.69 (0.42-0.95) | 0.74 (0.51-0.97) |
| ≥12 | 693 (85.6) | 0.74 (0.64-0.83) | 0.77 (0.67-0.87) |
| Age range (y) |  |  |  |
| 18-34 | 297 (36.6) | 0.74 (0.60-0.89) | 0.79 (0.65-0.93) |
| 35-59 | 419 (51.6) | 0.72 (0.59-0.84) | 0.76 (0.63-0.89) |
| ≥60 | 96 (11.8) | 0.71 (0.42-1.00) | 0.68 (0.39-0.98) |

Nova-WPF score, Nova score of whole plant foods; Nova-UPF score, Nova score of ultra-processed foods.

PABAK, Prevalence-Adjusted and Bias-Adjusted Kappa.
